# Enhanced Contact Investigations for Nine Early Travel-Related Cases of SARS-CoV-2 in the United States

**DOI:** 10.1101/2020.04.27.20081901

**Authors:** Rachel M. Burke, Sharon Balter, Emily Barnes, Vaughn Barry, Karri Bartlett, Karlyn D. Beer, Isaac Benowitz, Holly M. Biggs, Hollianne Bruce, Jonathan Bryant-Genevier, Jordan Cates, Kevin Chatham-Stephens, Nora Chea, Howard Chiou, Demian Christiansen, Victoria Chu, Shauna Clark, Sara Cody, Max Cohen, Erin E. Conners, Vishal Dasari, Patrick Dawson, Traci DeSalvo, Matthew Donahue, Alissa Dratch, Lindsey Duca, Jeffrey Duchin, Jonathan W. Dyal, Leora R. Feldstein, Marty Fenstersheib, Marc Fischer, Rebecca Fisher, Chelsea Foo, Brandi Freeman-Ponder, Alicia M. Fry, Jessica Gant, Romesh Gautom, Isaac Ghinai, Prabhu Gounder, Cheri T. Grigg, Jeffrey Gunzenhauser, Aron J. Hall, George S. Han, Thomas Haupt, Michelle Holshue, Jennifer Hunter, Mireille B. Ibrahim, Max W. Jacobs, M. Claire Jarashow, Kiran Joshi, Talar Kamali, Vance Kawakami, Moon Kim, Hannah Kirking, Amanda Kita-Yarbro, Rachel Klos, Miwako Kobayashi, Anna Kocharian, Misty Lang, Jennifer Layden, Eva Leidman, Scott Lindquist, Stephen Lindstrom, Ruth Link-Gelles, Mariel Marlow, Claire P. Mattison, Nancy McClung, Tristan D. McPherson, Lynn Mello, Claire M. Midgley, Shannon Novosad, Megan T. Patel, Kristen Pettrone, Satish K. Pillai, Ian W. Pray, Heather E. Reese, Heather Rhodes, Susan Robinson, Melissa Rolfes, Janell Routh, Rachel Rubin, Sarah L. Rudman, Denny Russell, Sarah Scott, Varun Shetty, Sarah E. Smith-Jeffcoat, Elizabeth A. Soda, Chris Spitters, Bryan Stierman, Rebecca Sunenshine, Dawn Terashita, Elizabeth Traub, Grace E. Vahey, Jennifer R. Verani, Megan Wallace, Matthew Westercamp, Jonathan Wortham, Amy Xie, Anna Yousaf, Matthew Zahn

## Abstract

**Background:** Coronavirus disease 2019 (COVID-19), the respiratory disease caused by the severe acute respiratory syndrome coronavirus 2 (SARS-CoV-2), was first identified in Wuhan, China and has since become pandemic. As part of initial response activities in the United States, enhanced contact investigations were conducted to enable early identification and isolation of additional cases and to learn more about risk factors for transmission.

**Methods:** Close contacts of nine early travel-related cases in the United States were identified. Close contacts meeting criteria for active monitoring were followed, and selected individuals were targeted for collection of additional exposure details and respiratory samples. Respiratory samples were tested for SARS-CoV-2 by real-time reverse transcription polymerase chain reaction (RT-PCR) at the Centers for Disease Control and Prevention.

**Results:** There were 404 close contacts who underwent active monitoring in the response jurisdictions; 338 had at least basic exposure data, of whom 159 had ≥1 set of respiratory samples collected and tested. Across all known close contacts under monitoring, two additional cases were identified; both secondary cases were in spouses of travel-associated case patients. The secondary attack rate among household members, all of whom had ≥1 respiratory sample tested, was 13% (95% CI: 4 – 38%).

**Conclusions:** The enhanced contact tracing investigations undertaken around nine early travel-related cases of COVID-19 in the United States identified two cases of secondary transmission, both spouses. Rapid detection and isolation of the travel-associated case patients, enabled by public awareness of COVID-19 among travelers from China, may have mitigated transmission risk among close contacts of these cases.

## Introduction

In late December 2019, a cluster of patients with pneumonia of unknown etiology was identified in Wuhan, China.^1^ This disease, now termed coronavirus disease 2019 (COVID-19), spread rapidly within China and quickly became pandemic.^1^ Phylogenetic analyses of severe acute respiratory syndrome coronavirus 2 (SARS-CoV-2), the causative agent of COVID-19, suggest zoonotic origin,^2^ with subsequent rapid spread indicative of person-to-person transmission.^3,4^ On January 20, 2020, the first case of COVID-19 was confirmed in the United States;^5^ through the end of January, nine more travel-related cases were identified. To interrupt transmission and facilitate early identification of secondary cases, public health authorities at the state, county, and local levels, in consultation with subject-matter experts from the U.S. Centers for Disease Control and Prevention (CDC), mobilized rapidly to place the patients under appropriate isolation and identify contacts exposed to these patients. Enhanced contact investigations were conducted centered on nine patients, and included in-depth interviews to better characterize exposure type, exposure duration, and contact medical history, in addition to collection of respiratory specimens to identify asymptomatic or pre-symptomatic infections. This report summarizes results from these nine enhanced contact investigations, including information on exposures, active monitoring, laboratory testing, and secondary attack rates among close contacts.

## Methods

### Contact Investigation Overview

These investigations included the close contacts of nine travel-related COVID-19 patients (hereafter referred to as “travel-associated case patients”) first identified as suspected COVID-19 patients between January 19 and January 30, 2020, and represent collaboration among state and local health department staff, CDC personnel, and healthcare facility staff (hereafter referred to as “COVID-19 Outbreak Response Teams”). The aims of these investigations were to interrupt transmission, investigate risk factors for transmission, and identify both symptomatic and asymptomatic infections among contacts of travel-associated case patients.

After diagnosis confirmation, these nine travel-associated case patients were interviewed to determine the date of symptom onset and to collect information on movements and interactions with other individuals during the presumed infectious period of the travel-associated case patient. Individuals with potential exposure to a travel-associated case patient were contacted by COVID-19 Outbreak Response Teams to determine their exposure risk level, whether they met the definition of a close contact, and whether they met criteria for active monitoring (terms defined below in “Classification of Contacts and Exposures”). Individuals not meeting the definition of a close contact are not included in this report.

To better understand SARS-CoV-2 transmission, COVID-19 Outbreak Response Teams selected a convenience sample of the close contacts who met active monitoring criteria to target for collection of additional, detailed exposure and demographic information using standardized forms; some sites were able to collect information on all close contacts, while others collected information only on those close contacts they classified as having had high-or medium-risk exposures. To understand the prevalence of asymptomatic or pre-symptomatic infection, a convenience sample of actively monitored close contacts was selected from whom to request respiratory (nasopharyngeal [NP] and oropharyngeal [OP]) samples outside of diagnostic specimen collection procedures (i.e., while contacts were asymptomatic or, in some cases, symptomatic with ≥ 1 previous negative SARS-CoV-2 result); some sites were able to request at least one set of samples from all close contacts, but most sites targeted sample collection mainly to close contacts determined to have had high-risk exposures, such as household members.

### Classification of Contacts and Exposures

Definitions of the infectious period, close contact, and exposure risk, as well as decisions regarding which types of contacts to monitor and how to manage their movement, were made at the site level by the COVID-19 Outbreak Response Teams. In one site, the presumed infectious period began on the date of symptom onset in the travel-associated case patient; however, in other sites, investigators also considered between one and three days before symptom onset in the travel-associated case patient. In all sites, the presumed infectious period ended when the travel-associated case patient was released from isolation. Generally, close contacts were defined as persons having frequent or more than brief contact (>1 – 2 minutes within 6 ft) with a travel-associated case patient during the travel-associated case patient’s presumed infectious period. Most sites also considered persons sharing the same airspace as the travel-associated case patient for more than ten minutes to be close contacts. For persons whose exposure occurred in the healthcare setting, the definition of close contact also depended on usage of Personal Protective Equipment (PPE) by the exposed person during their interaction with the travel-associated case patient.

Close contacts were grouped by type. Close contacts were classified as “Household contacts” if they were family members or friends of a travel-associated case patient who spent at least one night in the same residence as the travel-associated case patient during the presumed infectious period. Close contacts were classified as “Healthcare Personnel” (HCP) if they were healthcare or public health personnel working in healthcare settings who had the potential for exposure to a travel-associated case patient or to infectious materials from a travel-associated case patient. Close contacts who were defined as neither household contacts nor HCP were classified as “Community Contacts”; this category was broken down into contact occurring in a healthcare setting (e.g., waiting room close contacts) and contact occurring outside a healthcare setting (e.g., coworkers). “Community Contacts” also included flight-related close contacts of travel-associated case patients whose infectious periods included plane travel; these were defined as passengers seated in the same row as the travel-associated case patient, or in the two rows in front of or behind the travel-associated case patient.

For the purposes of analyses of HCP PPE data, “all recommended PPE for airborne and contact precautions” is defined as gloves, gown, eye protection (goggles or a disposable face shield that covers the front and sides of the face), and a Powered Air-Purifying Respirator (PAPR) or N95 filtering facepiece respirator (FFR) with fit-testing in the past year”; “all recommended PPE for droplet and contact precautions” is defined as gloves, gown, eye protection, and a face mask.^6^ Any encounter during which an HCP was wearing any combination of PPE not meeting either of the above definitions was considered to be “less protected.”

Risk stratification varied by site, but all sites considered household contacts as well as HCP who provided patient care without all recommended PPE to have had high-risk exposures.

### Data Collection

Close contacts meeting criteria for active monitoring were contacted daily via phone, text message, email, or in person, and were asked to report temperature and any symptoms. Close contacts selected for collection of detailed exposure information were interviewed using forms that were standardized within but not across jurisdictions. Information collected included dates of exposure to the travel-associated case patient; basic demographics; duration and types of exposure (e.g., face-to-face contact, direct physical contact); and, among HCP close contacts, use of PPE and interactions with or procedures performed on travel-associated case patients. Information was entered into standardized forms, spreadsheets, or databases maintained by the COVID-19 Outbreak Response Teams.

Close contacts selected for biological sample collection agreed to provide NP and OP samples for SARS-CoV-2 testing. Samples were collected by trained personnel. Most sites aimed to collect samples from close contacts with high-risk exposures every 2 – 3 days through 14 days following last exposure to the travel-associated case patient.

### Management of Close Contacts of COVID-19 Travel-Associated Case Patients

Close contacts living outside the response jurisdictions were forwarded to their respective health jurisdictions for assessment and monitoring. Some flight-related close contacts were forwarded to CDC’s Division of Migration and Quarantine for follow-up. Close contacts meeting criteria for active monitoring were monitored through 14 days following their last exposure to a travel-associated case patient. Close contacts not meeting criteria for active monitoring were asked to self-monitor and to contact the local health department if they developed any symptoms. Close contacts who developed new or worsening symptoms that COVID-19 Outbreak Response Teams deemed possibly consistent with COVID-19 (e.g., fever, cough) were considered to be suspect COVID-19 patients and had respiratory samples (and serum samples, when possible) collected and sent for testing at the time that determination was made, regardless of whether previous samples had been collected.

Household contacts of some non-hospitalized travel-associated case patients were quarantined in the same home with their respective travel-associated case patient following the travel-associated case patient’s diagnosis; this quarantine lasted through the period of case isolation. This situation is hereafter referred to as “co-habitation during case isolation.” To prevent household transmission, the travel-associated case patients were instructed to stay in a separate bedroom, use a separate bathroom if possible, and to wear a face mask if they had to be in the same room as cohabitants^7^.

### Laboratory Methods

All specimen samples were collected according to CDC guidance then packaged and shipped to CDC^8^. Specimens were refrigerated at 2–8°C before shipping on icepacks. CDC performed real-time reverse transcriptase-polymerase chain reaction (rRT-PCR) to detect three separate genetic markers of SARS-CoV-2 in the N1, N2, and N3 regions.^9^ Respiratory specimens were considered positive if all three genetic markers were positive by rRT-PCR.

### Statistical Methods and Data Management

Demographic, clinical, exposure, and testing results for contacts are summarized and described. Because the exact information that was collected varied by site, denominators are presented throughout. Collection of respiratory specimens is described by sample type and days since last exposure to the travel-associated case patient. The secondary attack rate by close contact type was calculated as the percent of close contacts with at least one positive SARS-CoV-2 test result among close contacts for whom ≥1 set of respiratory samples was tested; 95% confidence intervals were calculated using the Wilson Score interval. Differences in continuous variables were tested by the Mann-Whitney U test given non-parametric distributions.

Data were entered into Access, Excel, or REDCap databases. Data were merged and analyzed using the R Environment for Statistical Computing.

### Ethical Review

These activities were reviewed by CDC’s Human Research Protection Office and deemed to be non-human subjects research because they were conducted as part of a public health response.

## Results

### Description of the Close Contacts

A total of 553 (range 18 to 222 close contacts per case) individuals were identified as close contacts of the first nine patients with travel-related COVID-19 in the United States (**Figure 1**). Four hundred and four (73%; range: 1 to 206 per confirmed case) met criteria and participated in local active monitoring. Most of the 149 close contacts who did not participate in local active monitoring were determined to have had low-risk exposures that did not require active monitoring. Other close contacts who did not participate in local active monitoring were either unable to be reached or were managed by other jurisdictions.

**Figure 1:**
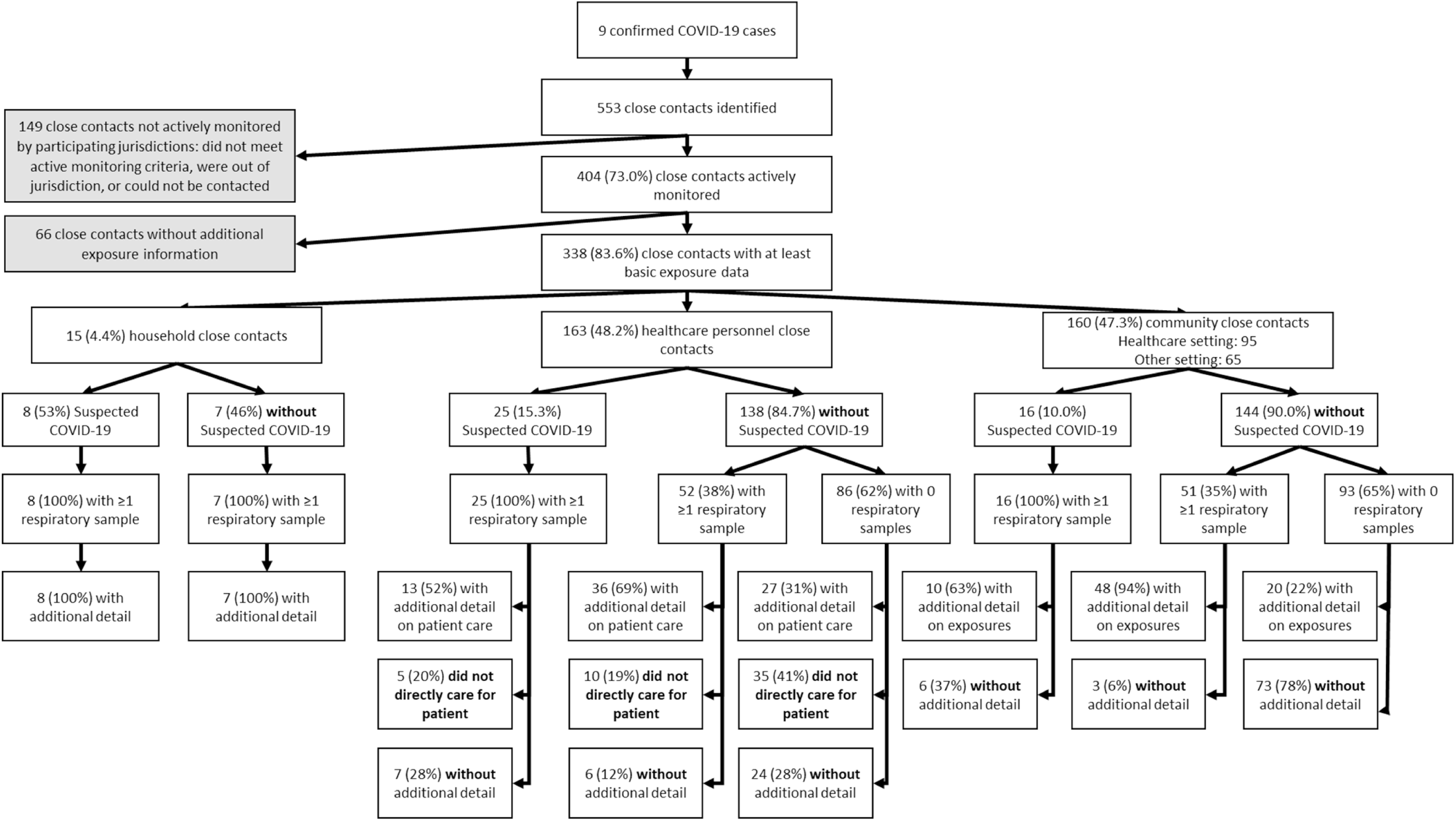
Flow Diagram of Close Contacts Identified and Included in Enhanced Contact Investigations Centered on 9 COVID-19 Patients, United States, January – February 2020

Among those close contacts actively monitored, 338 (84%) had either limited demographic and exposure type information (hereafter referred to as “at least basic exposure data”) or demographic plus detailed exposure information collected; the type and amount of information for different types of close contacts varied by study site depending on their protocol.

Of these 338 close contacts, 15 (4%) were household contacts, 163 (48%) were HCP, 95 (28%) were community contacts exposed in a healthcare setting, and 65 (19%) were community contacts exposed in another setting (**Table 1; Figure 1**). Among the actively monitored close contacts with demographic information, the majority were female (n=179/283; 63%) and aged 18–44 years (n=152/251; 61%). Of these 338 close contacts, only nine (3%) were exposed prior to symptom onset in the travel-associated case patient, including seven household members who were also exposed while the travel-associated case patient was symptomatic (data not shown).

**Table 1:** Demographic, Clinical, Exposure, and Investigation Characteristics Among Actively Monitored Close Contacts with at least Basic Data on Exposures to 9 Confirmed COVID-19 Patients, United States, January – February

|  | Household<br>N (%) | Healthcare<br>Personnel*<br>N (%) | Community:<br>Healthcare Setting<br>N (%) | Community:<br>Other Setting<br>N (%) |
| --- | --- | --- | --- | --- |
| *N* | 15 | 163 | 95 | 65 |
| **Demographics** |  |  |  |  |
| *Sex* |  |  |  |  |
| Female | 6 (40.0) | 106 (65.0) | 40 (42.1) | 27 (41.5) |
| Male | 9 (60.0) | 35 (21.5) | 28 (29.5) | 32 (49.2) |
| Missing Sex Information | 0 (0.0) | 22 (13.5) | 27 (28.4) | 6 (9.2) |
| *Age Category* |  |  |  |  |
| < 10 years | 1 (6.7) | 0 (0.0) | 6 (6.3) | 0 (0.0) |
| 10 – 17 years | 1 (6.7) | 0 (0.0) | 4 (4.2) | 0 (0.0) |
| 18 – 44 years | 7 (46.7) | 101 (62.0) | 24 (25.3) | 20 (30.8) |
| 45 – 64 years | 5 (33.3) | 40 (24.5) | 16 (16.8) | 12 (18.5) |
| 65+ years | 1 (6.7) | 2 (1.2) | 10 (10.5) | 1 (1.5) |
| Missing Age Information | 0 (0.0) | 20 (12.3) | 35 (36.8) | 32 (49.2) |
| *Race / Ethnicity* |  |  |  |  |
| White, Non-Hispanic | 0 (0.0) | 68 (41.7) | 22 (23.2) | 9 (13.8) |
| Black, Non-Hispanic | 0 (0.0) | 4 (2.5) | 0 (0.0) | 0 (0.0) |
| Hispanic / Latino | 0 (0.0) | 33 (20.2) | 12 (12.6) | 0 (0.0) |
| Asian | 7 (46.7) | 39 (23.9) | 8 (8.4) | 13 (20.0) |
| Other | 0 (0.0) | 2 (1.2) | 0 (0.0) | 0 (0.0) |
| Missing Race / Ethnicity Information | 8 (53.3) | 17 (10.4) | 53 (55.8) | 43 (66.2) |
| **Medical History** |  |  |  |  |
| *Smoking History* |  |  |  |  |
| Current | 3 (20.0) | 2 (1.2) | 2 (2.1) | 2 (3.1) |
| Former | 0 (0.0) | 10 (6.1) | 4 (4.2) | 3 (4.6) |
| Never | 4 (26.7) | 22 (13.5) | 15 (15.8) | 25 (38.5) |
| Missing Smoking Information | 8 (53.3) | 129 (79.1) | 74 (77.9) | 35 (53.8) |
| *Cardiovascular Disease* |  |  |  |  |
| No History | 4 (26.7) | 33 (20.2) | 13 (13.7) | 29 (44.6) |
| Yes | 1 (6.7) | 3 (1.8) | 9 (9.5) | 3 (4.6) |
| Missing Information | 10 (66.7) | 127 (77.9) | 73 (76.8) | 33 (50.8) |
| *Chronic Lung Disease* |  |  |  |  |
| No History | 5 (33.3) | 32 (19.6) | 17 (17.9) | 28 (43.1) |
| Yes | 1 (6.7) | 4 (2.5) | 5 (5.3) | 5 (7.7) |
| Missing Information | 9 (60.0) | 127 (77.9) | 73 (76.8) | 32 (49.2) |
| *Immunosuppressive Condition or Therapy\*\** |  |  |  |  |
| No History | 6 (40.0) | 39 (23.9) | 25 (26.3) | 33 (50.8) |
| Yes | 0 (0.0) | 1 (0.6) | 1 (1.1) | 0 (0.0) |
| Missing Information | 9 (60.0) | 123 (75.5) | 69 (72.6) | 32 (49.2) |

**Table 1: Demographic, Clinical, Exposure, and Investigation Characteristics Among Actively Monitored Close Contacts with at least Basic Data on Exposures to 9 Confirmed COVID-19 Patients, United States, January – February 2020**
|  | **Household**<br>**N (%)** | **Healthcare**<br>**Personnel\***<br>**N (%)** | **Community:**<br>**Healthcare Setting**<br>**N (%)** | **Community:**<br>**Other Setting**<br>**N (%)** |
| --- | --- | --- | --- | --- |
| *Diabetes Mellitus* |  |  |  |  |
| No History | 6 (40.0) | 29 (17.8) | 18 (18.9) | 30 (46.2) |
| Yes<sup>†</sup> | 0 (0.0) | 0 (0.0) | 4 (4.2) | 1 (1.5) |
| Missing Information | 9 (60.0) | 134 (82.2) | 73 (76.8) | 34 (52.3) |
| **Exposure Setting** |  |  |  |  |
| Private Home | 15 (100.0) | 2 (1.2) | 0 (0.0) | 1 (1.5) |
| Healthcare | 0 (0.0) | 161 (98.8) | 95 (100.0) | 0 (0.0) |
| Workplace (non-Healthcare) | 0 (0.0) | 0 (0.0) | 0 (0.0) | 25 (38.5) |
| Transit / Transit Hub | 0 (0.0) | 0 (0.0) | 0 (0.0) | 37 (56.9) |
| Other | 0 (0.0) | 0 (0.0) | 0 (0.0) | 2 (3.1) |
| **Investigation Procedures** |  |  |  |  |
| *Development of New or Worsening Symptoms (Suspected COVID-19)* |  |  |  |  |
| No | 7 (46.7) | 138 (84.7) | 87 (91.6) | 57 (87.7) |
| Yes | 8 (53.3) | 25 (15.3) | 8 (8.4) | 8 (12.3) |
| *Respiratory Sample Collection* |  |  |  |  |
| No samples | 0 (0.0) | 86 (52.8) | 72 (75.8) | 21 (32.3) |
| At least one set of samples | 15 (100.0) | 77 (47.2) | 23 (24.2) | 44 (67.7) |
| More than one set of samples | 15 (100.0) | 13 (8.0) | 3 (3.2) | 7 (10.8) |

\*Includes healthcare or public health professionals providing patient care or directly interacting with a COVID-19 patient in healthcare setting; cleaning personnel who potentially had contact with bodily fluids of a COVID-19 patient in a healthcare setting; and laboratory personnel who processed samples without recommended PPE. \*\*Contacts were asked about any non-cancer immunosuppressive conditions or therapies (e.g., therapy for autoimmune conditions such as rheumatoid arthritis), as well as about whether cancer chemotherapy had been received within the last 12 months.
<sup>†</sup>All persons reporting a history of diabetes specified type 2 diabetes.

A total of 49 actively monitored close contacts with at least basic exposure data (14%) developed new or worsening respiratory symptoms and had a set of diagnostic respiratory samples collected; 19 of these 49 (39%) had additional respiratory samples taken as part of enhanced contact investigations or case follow-up. There were 289 (86%) actively monitored close contacts with at least basic exposure data who remained without new or worsening symptoms, of whom 110 (38%) had at least one set of respiratory samples collected and 19 had more than one set of respiratory samples collected (**Figure 1**). A histogram of samples collected by type, contact category, and days since last exposure is shown in **Supplemental Figures 1 and 2**.

### Findings by Contact Type

#### Household Contacts

The nine travel-related travel-associated case patients had a total of 15 household contacts, of whom all were actively monitored. Two household contacts, both spouses of travel-associated case patients, became symptomatic and tested positive for SARS-CoV-2 (**Table 2**). The secondary attack rate among all household members of travel-associated case patients was 13% (95% CI: 4 – 38%). The secondary attack rate among significant others of travel-associated case patients was 25% (95% CI: 7 – 59%).

**Table 2:** Description of Household Contacts of Confirmed Travel-Acquired COVID-19 Case Patients (N = 15), United States, January – February

|  | N (%) | Any respiratory sample<br>N (row %) | >1 respiratory sample<br>N (row %) | Secondary cases detected<br>N (row %) |
| --- | --- | --- | --- | --- |
| *Overall* | 15 (100.0) | 15 (100.0) | 15 (100.0) | 2 (13.3) |
| *Relationship* |  |  |  |  |
| Significant Other | 8 (53.3) | 8 (100.0) | 8 (100.0) | 2 (25.0) |
| Other Household Member | 7 (46.7) | 7 (100.0) | 7 (100.0) | 0 (0.0) |
| *Exposure to Travel-Acquired COVID-19 Patient Following Diagnosis\** |  |  |  |  |
| Co-habiting with Case Patient During Case Isolation** | 5 (35.7) | 5 (100.0) | 5 (100.0) | 0 (0.0) |
| Isolated from Patient Following Diagnosis<sup>†</sup> | 9 (64.3) | 9 (100.0) | 9 (100.0) | 1 (11.1) |
| < 12 hours of exposure to the travel-associated case patient before isolation | 2 (22.2) | 2 (100.0) | 2 (100.0) | 0 (0.0) |
| 1 – 3 days of exposure to the travel-associated case patient before isolation | 5 (55.6) | 5 (100.0) | 5 (100.0) | 0 (0.0) |
| > 3 days of exposure to the travel-associated case patient before isolation | 2 (22.2) | 2 (100.0) | 2 (100.0) | 1 (50.0) |

\*Does not include 1 household member who was concurrently diagnosed following co-isolation with their spouse, who was a suspected case. \*\*These persons inhabited the same house as the travel-related COVID-19 patient throughout the period of case isolation, but were instructed to have the case patient stay in a separate bedroom and use a separate bathroom if possible, and to have the case patient wear a face mask if they could not avoid being in the same room. <sup>†</sup>These persons maintained separate living quarters from the travel-associated case patient throughout the period of case isolation.

There were two couples (“A” and “B”) in which secondary transmission occurred; neither spouse shared a travel history with their respective travel-associated case patient. In couple A, the travel-associated case patient developed symptoms one day after returning from travel and enrolled in active monitoring with the local health department; the spouse, who had no reported comorbidities, developed symptoms three days after the travel-associated case patient’s symptom onset. Couple A reported the use of some isolation precautions during the period of exposure: the travel-associated case patient wore a face mask and used a separate bedroom and bathroom even prior to developing symptoms, although the couple did share a meal and spent extended time in the car together. Both persons in Couple A were diagnosed with COVID-19 at the same time and were instructed to co-isolate at home until released by public health officials; however, eight days after symptom onset in the travel-associated case patient, couple A had to be hospitalized due to worsening conditions in both patients. In couple B, as in couple A, the travel-associated case patient’s symptoms began the day after returning from travel. Six days after symptom onset, the travel-associated case patient was hospitalized; the next day, the travel-associated case patient was identified as having suspect COVID-19, and the spouse entered active monitoring. Symptom onset in the couple B spouse could not be precisely determined, as this person has underlying conditions, but could have been as early as three days after the travel-associated case patient’s symptom onset. Couple B did not report any isolation precautions prior to the diagnosis of the travel-associated case patient (which occurred 8 days after symptom onset), and had frequent unmasked face-to-face and direct contact. In both couples, the spouse tested positive in the first respiratory samples collected.

In general, the length of exposure to the travel-associated case patient was lower among the 13 household contacts who did not develop COVID-19. Among these 13, eight (including 3 significant others) were completely separated from the travel-associated case patient once the patient was diagnosed and isolated; these persons maintained separate living space throughout the period of case isolation. Two (25%) had <12 hours of exposure to the symptomatic travel-associated case patient before isolation, five (63%) had 1 – 3 days of household exposure to the symptomatic travel-associated case patient before isolation, and one (13%) had eight days of household exposure to the symptomatic travel-associated case patient before isolation. Among these eight, three household members of two travel-associated case patients shared a travel history with the travel-associated case patient. All eight household members who were separated from the travel-associated case patients following diagnosis had serial specimens collected (median: 6.5 sets; range: 3 – 7), and all persistently tested negative; the date of last specimen collection ranged from five to 14 days following last exposure. Five household members (including 4 significant others) across four households co-habited with the travel-associated case patient during the period of case isolation; none shared a travel history with the travel-associated case patient. None of these travel-associated case patients were hospitalized at any time. When following up with these four co-habiting households via phone and in-person visits, the COVID-19 Outbreak Response Teams noted moderate to very high adherence to recommended home isolation procedures: in two out of four households, the travel-associated case patient was able to use a separate bathroom, and in only one of four households did the COVID-19 Outbreak Response Team ever directly observe or learn of the travel-associated case patient spending time in the same room as their household member(s). All five household members undergoing co-habitation had multiple respiratory samples collected for testing (median: 5 sets; range: 3 – 7) and remained SARS-CoV-2-negative throughout the period of co-habitation, with the last samples collected within seven days of the end of case isolation or the end of co-habitation. These samples are shown in **Supplemental Figure 3**.

#### Healthcare Personnel (HCP)

The nine travel-related travel-associated case patients made a total of 16 visits to healthcare facilities (3 outpatient clinics, 1 urgent care clinic, 12 hospital visits), resulting in 163 HCP contacts who met criteria for active monitoring. There were 126 HCP (77%) for whom data were available to describe patient contact and PPE usage in more detail. Among 49 HCP who provided care to or came into contact with the infectious fluids of travel-associated case patients and who had at least one set of respiratory samples collected and tested for SARS-CoV-2, the secondary attack rate was 0% (95% CI: 0 – 7%).

Two HCP (<1%) reported performing a possibly aerosol-generating procedure on a travel-associated case patient (nebulizer therapy: 1; spirometry: 1); both reported using all recommended PPE during the procedure. Laboratory personnel reported using recommended PPE when processing samples from travel-associated case patients.

Fourteen HCP (11%) reported collecting diagnostic respiratory specimens, of whom 6 (43%) reported always using all recommended PPE for airborne and contact precautions, 1 (7%) reported always using all recommended PPE for droplet and contact precautions, and 7 (50%) reported using less PPE than recommended on at least one occasion (i.e., PPE meeting neither airborne/contact nor droplet/contact precautions; **Table 3**). Among the 7 HCP using less PPE than recommended, 5 reported the details of their PPE use: all five wore gloves, one wore a gown, one wore eye protection, and one wore only a face mask; one wore an ill-fitting N95 FFR. All occasions during which “less-protected” HCP collected respiratory specimens from a travel-associated case patient were at the travel-associated case patient’s first presentation to care—before COVID-19 was confirmed.

**Table 3:** Exposures and Respiratory Sample Collection Among Actively Monitored Healthcare Personnel Providing Additional Information on Exposures and Use of Personal Protective Equipment (PPE) During Interactions with 9 COVID-19 Patients (N = 76), United States, January – February 2020

|  | N (%) | Any respiratory sample<br>N (row %)<sup>a</sup> | >1 respiratory sample<br>N (row %)<sup>a</sup> |
| --- | --- | --- | --- |
| **Collected Diagnostic Respiratory Specimens (N = 14)** |  |  |  |
| *PPE Category* |  |  |  |
| Airborne & Contact Precautions* | 6 (42.9) | 2 (33.3) | 0 (0.0) |
| Droplet & Contact Precautions** | 1 (7.1) | 1 (100.0) | 1 (100.0) |
| Did Not Meet Above PPE Definitions | 7 (50.0) | 5 (71.4) | 4 (57.1) |
| *Description of Missing PPE (N = 5)* |  |  |  |
| No Gloves | 0 (0.0) | N/A | N/A |
| No Gown | 4 (80.0) | 2 (50.0) | 1 (25.0) |
| No Eye Protection | 4 (80.0) | 2 (50.0) | 1 (25.0) |
| Respirator without Recent Fit Test | 1 (20.0) | 1 (100.0) | 1 (100.0) |
| Face Mask Only | 4 (80.0) | 2 (50.0) | 1 (25.0) |
| No Mask or Respirator | 0 (0.0) | N/A | N/A |
| **Provided Patient Care, Interacted with Patient, or Made Potential Contact with Infectious Fluids (N = 76)** |  |  |  |
| *PPE Category* |  |  |  |
| Airborne & Contact Precautions* | 33 (43.4) | 17 (51.5) | 0 (0.0) |
| Droplet & Contact Precautions** | 2 (2.6) | 0 (0.0) | 0 (0.0) |
| Did Not Meet Above PPE Definitions | 41 (53.9) | 32 (78.0) | 12 (29.3) |
| *Description of Missing PPE (N = 38)* |  |  |  |
| No Gloves | 9 (23.1) | 8 (88.9) | 4 (44.4) |
| No Gown | 28 (73.7) | 21 (75.0) | 6 (21.4) |
| No Eye Protection | 34 (89.5) | 26 (76.5) | 7 (20.6) |
| Respirator without Recent Fit Test | 2 (5.4) | 1 (50.0) | 1 (50.0) |
| Face Mask Only | 12 (30.8) | 8 (66.7) | 1 (8.3) |
| No Mask or Respirator | 13 (33.3) | 10 (76.9) | 3 (23.1) |

Abbreviations: N/A – not applicable; PPE – Personal Protective Equipment. <sup>a</sup>All respiratory samples tested negative for SARS-CoV-2. \*Defined as PAPR or N95 with fit test within last year, face shield / goggles, gloves, and gown. \*\*Defined as face mask, face shield / goggles, gloves, gown.

There were 76 (60%) HCP who reported providing patient care, touching the patient, or coming into contact with the patient’s infectious fluids (**Table 3**). Of these 76 HCP, 33 (43%) reported always using all recommended PPE for airborne and contact precautions, 2 (3%) reported always using all recommended PPE for droplet and contact precautions, and 41 (54%) reported using less PPE than recommended on at least one occasion. Among HCP using less PPE than recommended, 38 described PPE usage in detail. Eye protection was most frequently missing (34; 90%), followed by gown (28; 74%), and less commonly FFR or mask (13; 33%). All but three of these less-protected encounters occurred before COVID-19 was suspected in the travel-associated case patient. During all but seven of these less-protected encounters, the patient wore a mask for source control and removed it only for specimen collection and temperature measurement.

#### Community Contacts

There were 78 community contacts (24 exposed in a healthcare setting and 54 exposed in another setting) for whom information was sufficiently detailed to classify at least one specific type of exposure (**Table 4**). The secondary attack rate for community contacts of travel-associated case patients was 0% (95% CI: 0 – 6%) among the 58 contacts with some detailed exposure information and at least one set of respiratory samples tested.

**Table 4:** Exposures and Respiratory Sample Collection Among Actively Monitored Community Contacts Providing Additional Information on Exposure to 9 COVID-19 Patients (N = 78), United States, January – February 2020*

|  | Healthcare Setting |  |  | Other Setting |  |  |
| --- | --- | --- | --- | --- | --- | --- |
|  | N (%) | Any respiratory sample<br>N (row %)** | >1 respiratory sample<br>N (row %)** | N (%) | Any respiratory sample<br>N (row %)** | >1 respiratory sample<br>N (row %)** |
| *Face to Face Contact with Patient* |  |  |  |  |  |  |
| No | 18 (94.7) | 12 (66.7) | 1 (5.6) | 8 (22.9) | 6 (75.0) | 0 (0.0) |
| Yes | 1 (5.3) | 0 (0.0) | 0 (0.0) | 27 (77.1) | 20 (74.1) | 4 (14.8) |
| < 15 minutes | N/A | N/A | N/A | 11 (61.1) | 10 (100.0) | 1 (10.0) |
| ≥ 15 minutes | N/A | N/A | N/A | 7 (38.9) | 6 (85.7) | 2 (28.6) |
| *Direct Physical Contact with Patient* |  |  |  |  |  |  |
| No | 21 (100.0) | 14 (66.7) | 2 (9.5) | 26 (68.4) | 22 (84.6) | 4 (15.4) |
| Yes | 0 (0.0) | N/A | N/A | 12 (31.6) | 7 (58.3) | 0 (0.0) |
| *Was within 6 ft of Patient* |  |  |  |  |  |  |
| No | 7 (70.0) | 6 (85.7) | 1 (14.3) | 4 (10.5) | 4 (100.0) | 0 (0.0) |
| Yes | 3 (30.0) | 2 (66.7) | 1 (33.3) | 34 (89.5) | 31 (91.2) | 5 (14.7) |
| < 15 minutes | 3 (100.0) | 2 (66.7) | 1 (33.3) | 11 (57.9) | 9 (81.8) | 1 (9.1) |
| ≥ 15 minutes | 0 (0.0) | N/A | N/A | 8 (42.1) | 8 (100.0) | 3 (37.5) |
| *Was within 6 ft while Patient Coughed* |  |  |  |  |  |  |
| No | 11 (100.0) | 10 (90.9) | 2 (18.2) | 20 (71.4) | 18 (90.0) | 3 (15.0) |
| Yes | 0 (0.0) | N/A | N/A | 8 (28.6) | 6 (75.0) | 1 (12.5) |
| *Took an Object from Patient* |  |  |  |  |  |  |
| No | 20 (100.0) | 13 (65.0) | 2 (10.0) | 11 (37.9) | 8 (72.7) | 0 (0.0) |
| Yes | 0 (0.0) | N/A | N/A | 18 (62.1) | 17 (94.4) | 4 (22.2) |
| *Rode in the Same Vehicle, sitting within 6 ft of Patient* |  |  |  |  |  |  |
| No | 24 (100.0) | 16 (66.7) | 3 (12.5) | 32 (59.3) | 24 (75.0) | 2 (6.2) |
| Yes | 0 (0.0) | 0 (0.0) | 0 (0.0) | 22 (40.7) | 18 (81.8) | 3 (13.6) |
| < 1 hour | N/A | N/A | N/A | 2 (13.3) | 2 (100.0) | 0 (0.0) |
| ≥ 1 hour | N/A | N/A | N/A | 13 (86.7) | 12 (92.3) | 3 (23.1) |
| *Was in Same Room as Patient* |  |  |  |  |  |  |
| No | 6 (35.3) | 5 (83.3) | 0 (0.0) | 2 (4.4) | 2 (100.0) | 0 (0.0) |
| Yes | 11 (64.7) | 4 (36.4) | 1 (9.1) | 43 (95.6) | 32 (74.4) | 4 (9.3) |
| < 15 minutes | 2 (66.7) | 0 (0.0) | 0 (0.0) | 5 (23.8) | 3 (100.0) | 1 (33.3) |

|  | Healthcare Setting |  |  | Other Setting |  |  |
| --- | --- | --- | --- | --- | --- | --- |
|  | N (%) | Any respiratory sample<br>N (row %)** | >1 respiratory sample<br>N (row %)** | N (%) | Any respiratory sample<br>N (row %)** | >1 respiratory sample<br>N (row %)** |
| 15 min. - < 3 hours | 1 (33.3) | 1 (100.0) | 0 (0.0) | 6 (28.6) | 5 (100.0) | 2 (40.0) |
| ≥ 3 hours | 0 (0.0) | N/A | N/A | 10 (47.6) | 8 (80.0) | 0 (0.0) |

\*A total of 24 community contacts exposed in the health care setting and 55 exposed in other settings had information sufficiently detailed to classify at least one specific type of exposure. Sample sizes within each exposure do not sum to these totals because data was either not collected by the COVID-19 Outbreak Response Team or was not reported by the participant. \*\*Zero samples tested positive for SARS-CoV-2.

For community contacts exposed in the healthcare setting, some exposure information was missing as the patient was not always identified to these persons. Out of 19 who provided information, only one (5%) recalled having face-to-face contact with the travel-associated case patient. As compared to community contacts in the healthcare setting, community contacts exposed outside the healthcare setting had more interaction with the travel-associated case patient, and included 17 airport quarantine station screeners, 17 workplace contacts, 13 flight contacts, four rideshare drivers, and three friends. Of those with data, many reported having face-to-face contact (27/35; 77%) with the travel-associated case patient or spending time within 6 feet of the patient (34/38; 90%), and nearly all (43/45; 96%) could remember being in the same room as the travel-associated case patient. Fewer (8/28; 29%) reported being within 6 feet of the patient while the patient was coughing.

## Discussion

The enhanced contact tracing investigations undertaken by the COVID-19 Outbreak Response Teams around nine early travel-related COVID-19 cases in the United States resulted in identification of two transmissions from two travel-associated case patients to their spouses. We did not find evidence of secondary infection with SARS-CoV-2 among the other 47 actively monitored close contacts who developed symptoms suspicious for COVID-19 nor among the 110 close contacts who submitted respiratory specimens for virologic testing and who did not develop new symptoms during the 14-day active monitoring period.

The results of these investigations suggest that the risk of secondary COVID-19 infection is high among close household contacts of confirmed COVID-19 patients, especially significant others. Additional detail on one of these transmissions has been previously published.^10^ Overall, the average exposure durations among household contacts were longer among those who developed COVID-19 compared with those who did not. The higher transmission risk observed for household contacts is consistent with published research on other coronaviruses as well as SARS-CoV-2.^3,11,12^ Interestingly, we did not detect any SARS-CoV-2 infections among household members who were co-habiting with their confirmed SARS-CoV-2-positive household members during the period of case isolation. Since the majority of these co-habiting case patients and close contacts adhered very well to home isolation precautions, it is possible that careful adherence to isolation protocols mitigated the transmission risk to household members co-habiting with travel-associated case patients during the period of case isolation.

We did not detect any symptomatic secondary cases of COVID-19 among the 389 non-household contacts who completed active monitoring, including 77 HCP contacts and 67 community contacts from whom respiratory samples were tested for SARS-CoV-2. This finding is similar to findings from early contact investigations in Europe and Asia,^13–16^ but in contrast to other data suggesting a much higher secondary attack rate.^17^ It is possible that asymptomatic secondary cases could have developed in persons from whom respiratory samples were not collected,^18,19^ or that secondary cases could have developed without being detected in respiratory samples (for instance, if the timing of shedding did not align with the timing of sample collection);^11^ follow-up serology could help to rule out some of these possibilities. The low number of secondary cases could also be related to the quick isolation of most of these travel-associated case patients, as found in studies of MERS-CoV.^20,21^. Out of 41 HCP with higher-risk exposures to a travel-associated case patient, all but three reported these exposures to have occurred prior to confirmation of the patient’s diagnosis. Once the patients were classified as suspected COVID-19 cases, contact and transmission-based precautions were begun and maintained by the majority of HCP throughout the patient’s care. In contrast to the first community-acquired cases identified in the United States, it is also important to consider that most if not all of these first travel-related cases were likely cognizant of the possibility that they were infected, and engaged in practices such as wearing face masks, practicing social distancing in some cases, and seeking care;^5,10,22^ these practices have the potential to have reduced secondary transmission from these cases. Further, these investigations were taking place at a very early stage in the U.S. outbreak, and many sites employed a low threshold for defining close contacts, potentially inflating our denominator.

This report is subject to several limitations. Because individual COVID-19 Outbreak Response Teams were operating out of distinct locations with unique considerations, the procedures of the investigation were heterogeneous across sites: different teams made different decisions regarding how to define close contacts, how to categorize exposure risk, which close contacts to actively monitor, which types of exposure information to collect, and how often (and from whom) to attempt collection of respiratory samples. Although available information was standardized across sites for the purposes of analysis, this heterogeneity led to missing data, limiting our ability to assess certain types of exposure in this population. Relatedly, only 47% of actively monitored close contacts provided any respiratory specimens, and fewer provided serial respiratory specimens. Incomplete sample collection could have biased secondary attack rates, and also raises the possibility of a missed asymptomatic infection. Symptomatic infections occurring after the end of the 14-day monitoring period also could have been missed. Although some sites attempted to reach individuals with close contact prior to symptom onset in the travel-associated case patient, many of these exposures occurred during the travel of the travel-associated case patient and therefore were managed by other jurisdictions. As a result, only nine such persons are included in the present analysis (of whom seven were household members with continued exposure). Thus, secondary transmissions occurring prior to symptom onset in the travel-associated case patient may have been missed. Missed transmissions (either asymptomatic infections, transmissions occurring outside the presumed infectious period, or transmissions with an incubation period outside the 14-day active monitoring period) may have contributed to community spread, as suggested by some genetic analysis of SARS-CoV-2 circulating in Washington.^23^ Lastly, although all sites endeavored to interview and sample the close contacts with the highest-risk exposures, the selection of the sample for interview and respiratory specimen collection was not systematic or consistent across sites. This may have introduced bias in the characteristics of the close contacts described as well as in the secondary attack rates.

The findings of these enhanced contact investigations, consistent with other research, suggest that household members of COVID-19 patients, particularly significant others, who are likely to have prolonged close contact with the patients, are at excess risk of developing COVID-19.^3,12^ Further, the lack of observed transmission to HCP underlines the importance and effectiveness of rapid identification and isolation of suspected cases of COVID-19 and appropriate use of PPE in caring for these patients^24,25^. Careful adherence to infection prevention procedures within the home and the healthcare setting may help to reduce the risk of transmission of COVID-19, while monitoring and quarantine of household contacts may help interrupt transmission. Close contacts with high-risk exposures (such as household contacts or HCP providing patient care without PPE) should continue to be quarantined and actively monitored, as data from COVID-19 clusters have raised the concern of transmission from asymptomatic or pre-symptomatic individuals infected with SARS-CoV-2,^18,19,26,27^ which may be contributing to the widespread community transmission that is currently occurring in the United States.^28^ It is important for HCP and public health professionals to remain vigilant to the possibility of SARS-CoV-2 transmission from confirmed cases to household, community, and HCP contacts, and continue to emphasize isolation and other precautions.^6,7^ Further, individuals exposed to SARS-CoV-2 should self-quarantine, and all persons should practice everyday behaviors to limit the spread of COVID-19 such as proper hand hygiene, covering coughs and sneezes, wearing cloth face coverings, and social distancing.^29,30^

## Data Availability

Data may be available upon reasonable request.

## ACKNOWLEDGMENTS

The authors would first like to acknowledge the COVID-19 patients and close contacts described in this manuscript. We also gratefully acknowledge the assistance and contributions of Glen Abedi, Sharad Aggarwal, Olivia Almendares, Alison Binder, Catherine Bozio, Nakia Clemmons, Aaron Curns, Rebecca Dahl, Connor Hoff, Michelle Kautz, Marie Killerby, Lindsay Kim, Stephanie Kujawski, Steven Langerman, Jessica Leung, Allison Miller, Christine Miner, Ruth N. Moro, Florence Whitehill, and Rebecca Woodruff of CDC; and Meredith Haddix of Santa Clara County Public Health.

## SUPPLEMENTAL MATERIAL

**Supplemental Figure 1:**
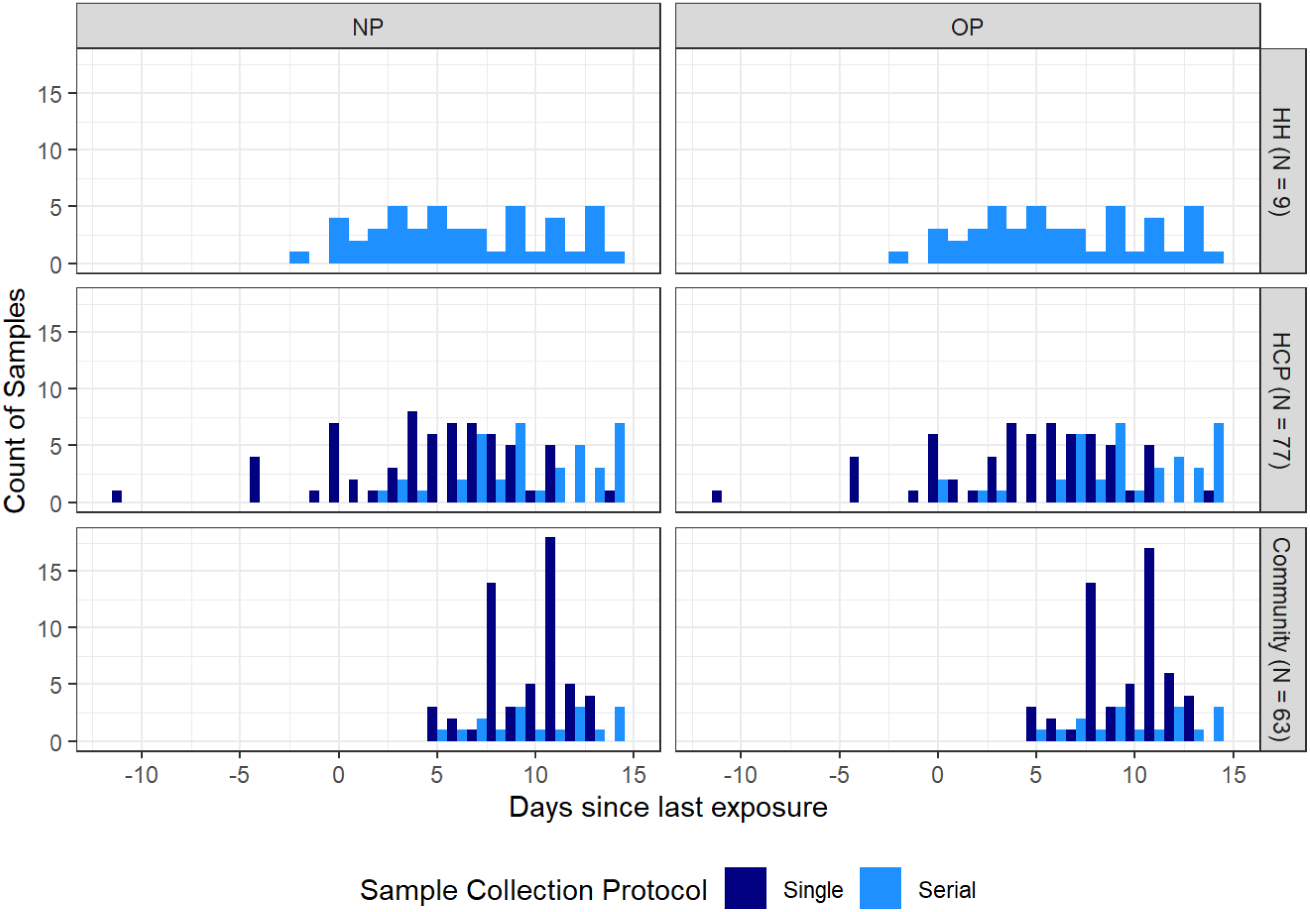
Histogram of Specimens Collected by Day Since Last Exposure, Grouped by Type of Contact and Days per Person. Specimens (N = 448) collected from 149 contacts with known exposure periods are shown by the days following last exposure to the confirmed COVID-19 patient on the x axis. Excludes specimens from five household contacts with ongoing household exposure after the diagnosis of the travel-associated case patient, one household member diagnosed concurrently as the travel-associated case patient, and four community contacts without a known exposure period or date of collection. Specimens from contacts submitting only a single set of respiratory specimens are shown in navy, while specimens from those submitting multiple sets of respiratory specimens are shown in lighter blue. Household contacts (HH) are shown in the first row (excluding 5 who were co-habiting with the case patient following diagnosis and 1 who was concurrently diagnosed), Healthcare Personnel (HCP) contacts are shown in the second row, and community contacts are shown in the bottom row. The first column shows the number of nasopharyngeal (NP) swabs collected and tested, and the second column shows the number of oropharyngeal (OP) swabs collected and tested. Some HH members and HCP have specimens collected at negative days from last exposure—these represent specimens collected during a period of ongoing exposure to the COVID-19 patient. Samples collected relative to the first day of exposure are shown in Supplemental Figure 1. Only the initial specimens are shown for the secondary cases, as these specimens tested positive.

**Supplemental Figure 2:**
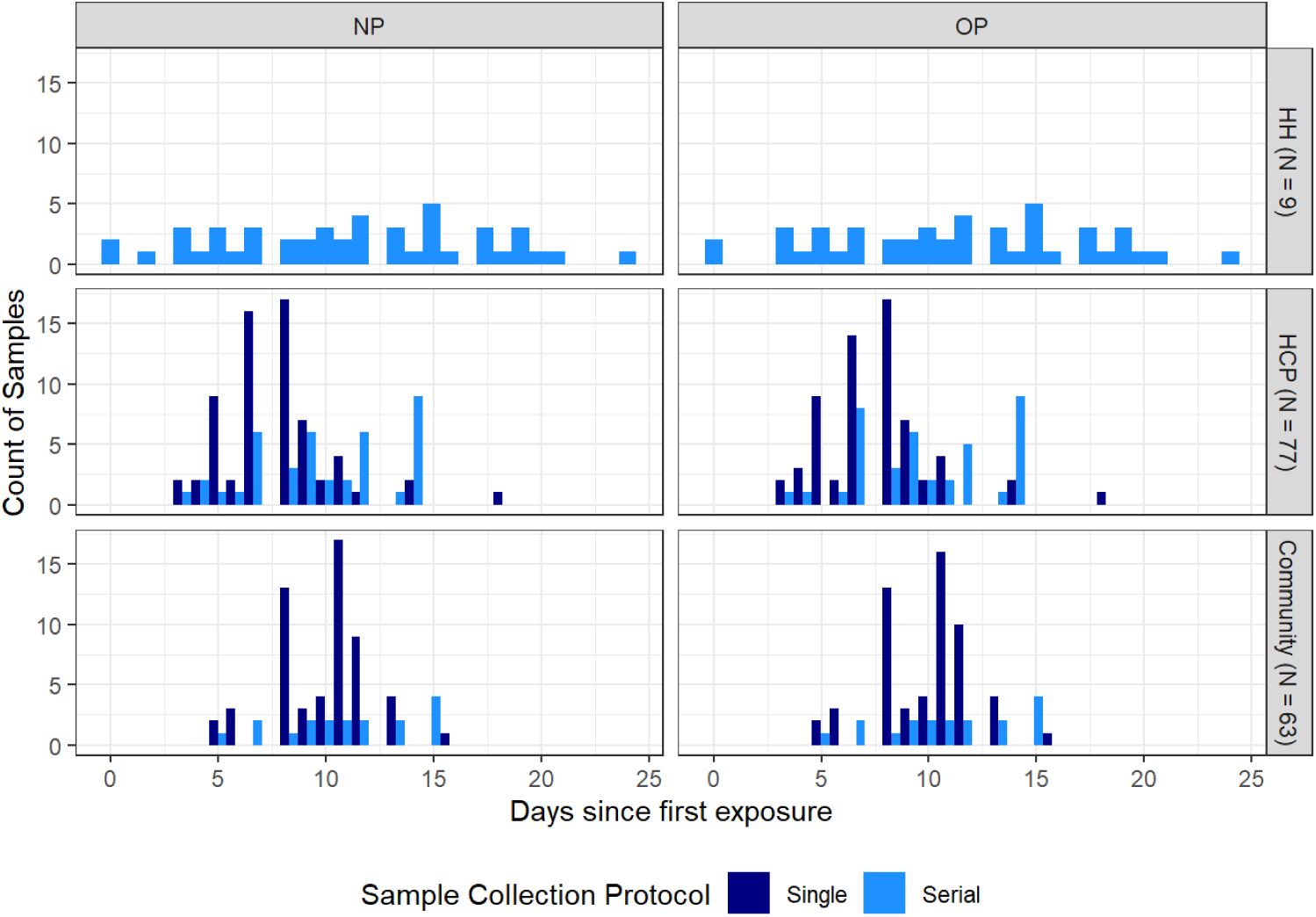
Histogram of Specimens Collected by Day Since First Exposure, Grouped by Type of Contact and Days per Person. Specimens (N = 448) collected from 149 contacts with known exposure periods are shown by the days following first exposure to the confirmed COVID-19 patient on the x axis. Excludes specimens from five household contacts with ongoing household exposure after the diagnosis of the travel-associated case patient, one household member diagnosed concurrently as the travel-associated case patient, and four community contacts without a known exposure period or date of collection. Specimens from contacts submitting only a single set of respiratory specimens are shown in navy, while specimens from those submitting multiple sets of respiratory specimens are shown in lighter blue. Household contacts (HH) are shown in the first row, Healthcare Personnel (HCP) contacts are shown in the second row, and community contacts are shown in the bottom row. The first column shows the number of nasopharyngeal (NP) swabs collected and tested, and the second column shows the number of oropharyngeal (OP) swabs collected and tested. Only the initial specimens are shown for the secondary cases, as these specimens tested positive.

**Supplemental Figure 3:**
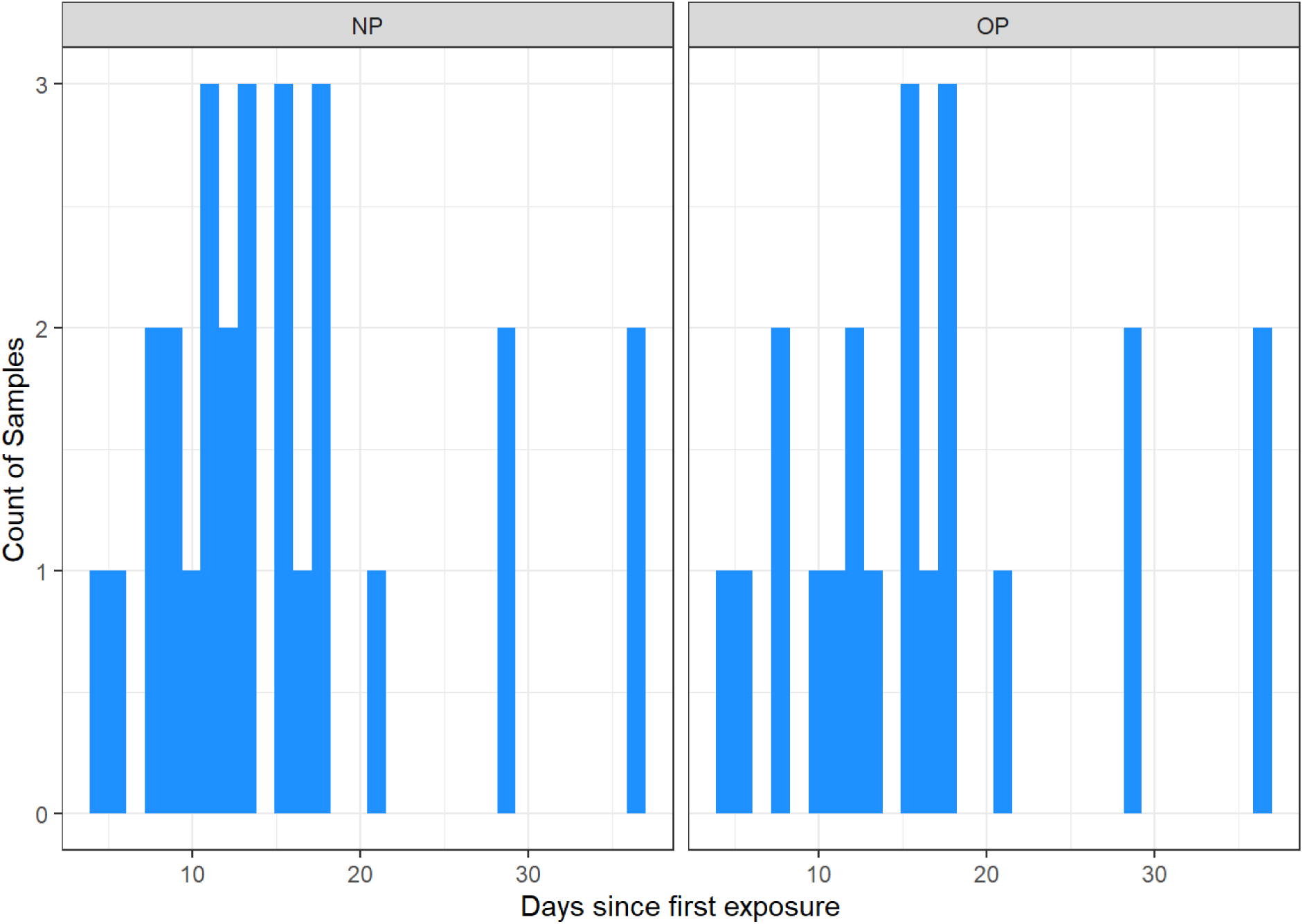
Histogram of Specimens Collected by Day Since First Exposure, Among Household Contacts Who Were Co-Habiting with Travel-Associated COVID-19 Case Patients During the Period of Case Isolation. Specimens (N = 48) collected from 5 contacts who were co-habiting with a COVID-19 patient following the travel-associated case patient’s diagnosis are shown by the days following first exposure to the confirmed COVID-19 patient on the x axis. The first column shows the number of nasopharyngeal (NP) swabs collected and tested, and the second column shows the number of oropharyngeal (OP) swabs collected and tested.

## REFERENCES

1. Coronavirus disease 2019 (COVID-19): Situation Report – 36. Geneva, Switzerland: World Health Organization, 2020.

2. Zhou P, Yang XL, Wang XG, et al. A pneumonia outbreak associated with a new coronavirus of probable bat origin. Nature 2020; 579(7798): 270–3.

3. Chan JF-W, Yuan S, Kok K-H, et al. A familial cluster of pneumonia associated with the 2019 novel coronavirus indicating person-to-person transmission: a study of a family cluster. The Lancet 2020; 395(10223): 514–23.

4. Li Q, Guan X, Wu P, et al. Early Transmission Dynamics in Wuhan, China, of Novel Coronavirus-Infected Pneumonia. N Engl J Med 2020.

5. Holshue ML, DeBolt C, Lindquist S, et al. First Case of 2019 Novel Coronavirus in the United States. N Engl J Med 2020.

6. Interim Infection Prevention and Control Recommendations for Patients with Suspected or Confirmed Coronavirus Disease 2019 (COVID-19) in Healthcare Settings. 1 April 2020 2020. https://www.cdc.gov/coronavirus/2019-ncov/hcp/infection-control-recommendations.html (accessed 5 April 2020).

7. If You Are Sick or Caring for Someone. https://www.cdc.gov/coronavirus/2019-ncov/if-you-are-sick/index.html (accessed 5 April 2020).

8. Interim guidelines for collecting, handling, and testing clinical specimens from persons under investigation (PUIs) for 2019 novel coronavirus (2019-nCoV). https://www.cdc.gov/coronavirus/2019-ncov/ lab/guidelines-clinical-specimens.html (accessed 10 March 2020).

9. Research Use Only 2019-Novel Coronavirus (2019-nCoV) Real-time RT-PCR Primer and Probe Information. April 10, 2020. https://www.cdc.gov/coronavirus/2019-ncov/lab/rt-pcr-panel-primer-probes.html (accessed 15 April 2020 2020).

10. Ghinai I, McPherson TD, Hunter JC, et al. First known person-to-person transmission of severe acute respiratory syndrome coronavirus 2 (SARS-CoV-2) in the USA. Lancet 2020.

11. Arwady MA, Alraddadi B, Basler C, et al. Middle East Respiratory Syndrome Coronavirus Transmission in Extended Family, Saudi Arabia, 2014. Emerg Infect Dis 2016; 22(8): 1395–402;.

12. Covid-19 National Emergency Response Center E, Case Management Team KCfDC, Prevention. Early Epidemiological and Clinical Characteristics of 28 Cases of Coronavirus Disease in South Korea. Osong Public Health Res Perspect 2020; 11(1): 8–14.

13. Covid-19 National Emergency Response Center E, Case Management Team KCfDC, Prevention. Coronavirus Disease-19: Summary of 2,370 Contact Investigations of the First 30 Cases in the Republic of Korea. Osong Public Health Res Perspect 2020; 11(2): 81–4.

14. Pung R, Chiew CJ, Young BE, et al. Investigation of three clusters of COVID-19 in Singapore: implications for surveillance and response measures. Lancet 2020; 395(10229): 1039–46.

15. Bernard Stoecklin S, Rolland P, Silue Y, et al. First cases of coronavirus disease 2019 (COVID-19) in France: surveillance, investigations and control measures, January 2020. Euro Surveill 2020; 25(6).

16. Haveri A, Smura T, Kuivanen S, et al. Serological and molecular findings during SARS-CoV-2 infection: the first case study in Finland, January to February 2020. Euro Surveill 2020; 25(11).

17. Liu Y, Eggo RM, Kucharski AJ. Secondary attack rate and superspreading events for SARS-CoV-2. Lancet 2020; 395(10227): e47.

18. Bai Y, Yao L, Wei T, et al. Presumed Asymptomatic Carrier Transmission of COVID-19. JAMA 2020.

19. Kimball A HK, Arons M, et al. Asymptomatic and Presymptomatic SARS-CoV-2 Infections in Residents of a Long-Term Care Skilled Nursing Facility — King County, Washington, March 2020.. MMWR Morb Mortal Wkly Rep 2020; (ePub: 27 March 2020.).

20. Kang CK, Song KH, Choe PG, et al. Clinical and Epidemiologic Characteristics of Spreaders of Middle East Respiratory Syndrome Coronavirus during the 2015 Outbreak in Korea. J Korean Med Sci 2017; 32(5): 744–9.

21. Hunter JC, Nguyen D, Aden B, et al. Transmission of Middle East Respiratory Syndrome Coronavirus Infections in Healthcare Settings, Abu Dhabi. Emerg Infect Dis 2016; 22(4): 647–56.

22. Patel A, Jernigan DB, nCo VCDCRT. Initial Public Health Response and Interim Clinical Guidance for the 2019 Novel Coronavirus Outbreak -United States, December 31, 2019-February 4, 2020. MMWR Morb Mortal Wkly Rep 2020; 69(5): 140–6.

23. Trevor Bedford, Alexander L. Greninger, Pavitra Roychoudhury, et al. Cryptic transmission of SARSCoV-2 in Washington State. *MedrXive*; 2020.

24. Cheng VCC, Wong SC, Chen JHK, et al. Escalating infection control response to the rapidly evolving epidemiology of the coronavirus disease 2019 (COVID-19) due to SARS-CoV-2 in Hong Kong. Infect Control Hosp Epidemiol 2020: 1–6.

25. Wong SC, Kwong RT, Wu TC, et al. Risk of nosocomial transmission of coronavirus disease 2019: an experience in a general ward setting in Hong Kong. J Hosp Infect 2020.

26. Moriarty LF, Plucinski MM, Marston BJ, et al. Public Health Responses to COVID-19 Outbreaks on Cruise Ships -Worldwide, February-March 2020. MMWR Morb Mortal Wkly Rep 2020; 69(12): 347–52;.

27. Wei WE LZ, Chiew CJ, Yong SE, Toh MP, Lee VJ. Presymptomatic Transmission of SARS-CoV-2 — Singapore, January 23–March 16, 2020. MMWR Morb Mortal Wkly Rep 2020.

28. Situation Summary. 2020. https://www.cdc.gov/coronavirus/2019-ncov/cases-updates/summary.html (accessed 20 March 2020).

29. How to Protect Yourself & Others. 2020. https://www.cdc.gov/coronavirus/2019-ncov/prevent-getting-sick/prevention.html (accessed 9 April 2020).

30. Social Distancing, Quarantine, and Isolation. 2020. https://www.cdc.gov/coronavirus/2019-ncov/prevent-getting-sick/social-distancing.html (accessed 9 April 2020).

